# The Delta Variant Had Negligible Impact on COVID-19 Vaccine Effectiveness in the USA

**DOI:** 10.1101/2021.09.18.21263783

**Authors:** Ben Blaiszik, Carlo Graziani, James L. Olds, Ian Foster

## Abstract

The SARS-CoV-2 Delta variant (B.1.617.2) was initially identified in India in December 2020. Due to its high transmissibility, its prevalence in the U.S.A. grew from a near-zero baseline in early May 2021 to nearly 100% by late August 2021, according to CDC tracking. We accessed openly available data sources from the public health authorities of seven U.S. states, five U.S. counties, and the District of Columbia on RT-PCR COVID-19 tests split by the COVID-19 vaccination status of individuals tested during this period. Together, these time series enable estimation and tracking of COVID-19 vaccine effectiveness (VE^∗^) (against RT-PCR diagnosed infection) concurrently with the growth of Delta variant prevalence in those locations. Our analyses reveal that in each locality the VE^∗^ for the combined set of all three US vaccines remained relatively stable and quite well-performing, despite the dramatic concurrent rise of Delta variant prevalence. We conclude that the Delta variant does not significantly evade vaccine-induced immunity. The variations in our measured VE^∗^ appear to be driven by demographic factors affecting the composition of the vaccinated cohorts, particularly as pertains to age distribution. We report that the measured VE^∗^, aggregated across the collected sites, began at a value of about 0.9 in mid-May, declined to about 0.76 by mid-July, and recovered to about 0.9 by mid-September.

**Summary:** We estimated local COVID-19 vaccine effectiveness using RT-PCR COVID-19 test data broken out by vaccination status from select localities in the U.S.A. between 15 May and 15 September 2021 while the SARS-CoV-2 Delta variant (B.1.617.2) was ascending from essentially zero prevalence to total dominance of the genome, and showed that the rise of the Delta variant had negligible effect on vaccine effectiveness.

## Introduction

In the first quarter of 2020, SARS-CoV-2, a novel beta coronavirus, disrupted global society by triggering a pandemic that, within a month of onset, locked down activity worldwide. That pandemic continues to the present (September 2021). The virus emulates its ancestor, SARS-CoV, by eliciting a high-affinity binding event between its spike envelope protein (S) and the human angiotensin converting enzyme II (ACE2) that allows it to gain entry into host epithelial cells in the host respiratory system and subsequently into other organs [1]. In contrast to SARS-CoV, SARS-CoV-2 is far more transmissible between its human hosts because its S protein binds to ACE2 with a much higher affinity [2]. The resultant disease, COVID-19, produces in patients a constellation of symptoms that vary widely but can result in hospitalization or death. As of September 2021, 219M diagnosed cases had resulted in 4.5M reported deaths globally.

The heterogeneity in clinical outcomes for COVID-19 early in the pandemic (before the advent of vaccines) is partly a function of the host’s innate [3] and adaptive immune system response [4]. There is a natural variance in phenotype for the various constituents of these systems. Even for immunologically-naive human hosts (as was the case for most of Earth’s human population in early 2020), there is some evidence for immunological cross-reactivity to SARS-CoV2 as a result of previous coronavirus exposure [5], leading to speculation that prior exposure to other coronaviruses might explain some of the phenotypic variance.

Another major factor in determining clinical outcomes is viral load [6]. In the case of non-vaccinated COVID-19 patients, an increased viral load would increase the frequency for viral RNA to enter host cells simply because there are more S-ACE2 binding events. Such logic has led to the design of engineered ACE2 to “divert” viral S [7] from its cellular target. In the case of vaccinated “breakthrough” patients, high viral load could overwhelm the initial innate immune response such that vaccine-induced neutralizing antibodies and other later-activated components of the adaptive system are responding to a viral infection that is already well-established.

All of these factors have acted together to create a challenge for policy makers and to create confusion among the public about COVID-19’s threat to human health [8].

### Vaccines, Variants, and “Breakthrough” Infections

National governments, often in partnership with the private sector, responded to the pandemic with an unprecedented program to rapidly develop vaccines, most directed against the S protein. In the United States, three vaccines were subsequently approved for emergency use by the FDA based on the results of phase‘3 clinical trials: two employing mRNA technology (Moderna [9, 10] and Pfizer/BioNTech [11, 12]) and one employing a modified adenovirus (Janssen JNJ-78436735 or Ad26.COV2.S) [13, 14]. To date, these vaccines have been administered to approximately 51% of the US population [15]. All three vaccines have proven to be effective in protecting fully vaccinated individuals from serious clinical complications of COVID-19.

At the beginning of the pandemic, it was believed that the virus mutation rate was too slow to be a concern [16], but this view has since been altered by the emergence of new variant strains, aided by some fitness advantage gained by mutation. In general, the molecular substrate for these fitness advantages is in the binding interface between the viral S and human ACE2 [17]. Among the variant strains, a few are considered of greater public health concern because of mutations that produce greater transmissibility, more severe disease, or decreased neutralization by human immune response, including by vaccine-induced immunity. In all such cases, viral load is a common factor affecting clinical outcomes [6].

Four such “variants of concern” have emerged to-date: the B.1.1.7 lineage (also called 20I/501Y.V1 or variant of concern [VOC] 202012/01), identified in the U.K. in December 2020 [18]; the B.1.351 lineage (20H/501Y.V2), first identified in the Republic of South Africa in December 2020 [19]; the P.1 lineage (20J/501Y.V3), identified in December 2020 in travelers from Brazil [20]; and the B.1.617.2 (Delta) variant identified in India in April 2021 [21].

In the current work, we address the concerns about infections reported in vaccinated individuals, colloquially described as “breakthrough infections” [22], in the context of the recent dominance of the Delta variant. The authors of [22] note that “Despite the high level of vaccine efficacy, a small percentage of fully vaccinated persons (i.e., those who have received all recommended doses of an FDA-emergency or fully authorized COVID-19 vaccine) will develop symptomatic or asymptomatic infections with SARS-CoV-2, the virus that causes COVID-19.” That is, breakthrough infections are expected from exposure to *any* strain of the virus, simply due to the fact that the protective efficacy and effectiveness of the vaccines is not 100%. This is a result of the immunological heterogeneity in phenotype described above, which leads to some individuals being more weakly stimulated by vaccines than others, and thus more likely to become infected upon exposure despite vaccination. Thus, some individuals are more weakly stimulated by vaccines than others, and more likely to become infected upon exposure despite vaccination. Hence, the real question about breakthrough infections with respect to Delta variant is not whether it causes them, but rather “How many more breakthrough infections per infectious exposure occur in consequence of the advent of the Delta variant than would have occurred otherwise?”.

### Data and Vaccine Effectiveness

We have found that in the U.S.A., the data required to explore the relationship between the Delta variant and vaccine effectiveness exist and are public, although they are not easy to find and are not collected in one place by one authority. The required data are aggregated reverse transcription–polymerase chain reaction (RT-PCR) test data, broken out by subject vaccination status, and reported at a regular cadence: preferably daily, and no less frequently than weekly. A few U.S. county or state health department aggregate their test data in this way. Thus, we have attempted to capture time series of data reported from the reporting sites we have uncovered. The sites are heterogeneous, both geographically and with respect to vaccination coverage. These sources are the public health departments of seven U.S. states, five U.S. counties, and the District of Columbia (Washington D.C.). Their details are listed in Table 1.

**Table 1:** Web resources on COVID vaccine breakthrough infections in the U.S.A. considered in this study. Collection methods: (MC) manual collection, (AC) automated collection, (S) image snapshot. N is the number of data samples. All dates are in 2021. All data sources are provided and described in the publicly available GitHub repository [23].

| Location | Type | Collection Method | Population | N | Date (mm-dd) |  |
| --- | --- | --- | --- | --- | --- | --- |
|  |  |  |  |  | From | To |
| Washington, DC | District of Columbia | MC | 705,749 | 11 | 06-14 | 08-30 |
| Contra Costa County, CA | County | MC | 1,159,507 | 17 | 05-13 | 09-09 |
| DuPage County, IL | County | S | 922,921 | 9 | 07-14 | 09-07 |
| King County, WA | County | MC | 2,269,675 | 15 | 05-26 | 09-08 |
| San Diego County, CA | County | S | 3,338,330 | 2 | 06-28 | 08-24 |
| Santa Clara County, CA | County | MC | 1,936,259 | 16 | 05-14 | 09-03 |
| Connecticut | State | S | 3,605,944 | 2 | 07-01 | 08-18 |
| Massachusetts | State | S | 7,033,469 | 3 | 07-30 | 08-21 |
| New York | State | MC | 13,877,519 | 3 | 06-30 | 07-19 |
| Oregon | State | S | 4,237,256 | 7 | 07-17 | 09-04 |
| South Carolina | State | S | 5,124,712 | 2 | 06-01 | 07-31 |
| Utah | State | AC | 3,310,774 | 17 | 05-14 | 09-10 |
| Virginia | State | MC | 8,631,393 | 17 | 05-15 | 09-11 |

With such time-series of test data, it is possible to estimate a measure of *vaccine effectiveness* (VE^∗^). This is a measure of vaccine performance that is distinct from vaccine efficacy (VE). VE is measured in the course of controlled clinical trials [24] of nearly identical treatment and placebo groups, and is therefore as entirely a clinical property of the vaccine itself as the trial design can attain. VE^∗^, on the other hand, is measured in the real world using unequal, potentially poorly-controlled groups of vaccinated and unvaccinated populations which evolve over time, and as such is a measure of real-world risk reduction—conditioned on a locality and time—due to vaccination.

The advantage of estimating VE^∗^ from testing data is that the aggregate results are a *time series* of VE^∗^, which show the evolving behavior of effectiveness per location and on average, and which can be compared to the time-series of other quantities, such as the growth of prevalence of the Delta strain. As we will see below, such a comparison is visually very instructive about the impact of the Delta variant on vaccine performance. The reason is that the rapid spread of Delta during the summer of 2021 provides a natural experiment. If vaccines are less effective against Delta, then as the Delta variant spread through the summer, we should expect to see an increased rate of breakthrough infections per infectious exposure, which is to say, a steady decrease in VE^∗^. On the other hand, if vaccine effectiveness remains unchanged, VE^∗^ should not systematically decrease.

We analyze the results from this natural experiment in the Results section, below. To anticipate the outcome, the answer to the question posed above is: *There is no evidence that the advent of the Delta variant has caused any more “breakthrough” infections per infectious exposure to occur than would have occurred had it never arrived in the U*.*S*.*A*.

There have been a number of well-controlled previous studies that were designed to assess the extent to which the effectiveness of COVID-19 vaccines is decreased by exposure to the Delta variant. These include studies showing moderate reductions in effectiveness in comparisons with the Alpha (B.1.1.7) variant in the U.K [25], Scotland [26], and in Canada [27]. A study in Qatar against Delta only [28] appeared to show diverging effectivenesses, higher for the Moderna vaccine and lower (with greater uncertainty) for the Pfizer vaccine, and some evidence for waning protection from the latter. In the U.S.A, a CDC study following health care personnel over time [29] appeared to show a substantial decrease of vaccine effectiveness (91% to 66%, albeit with large uncertainties) concurrently with the advent of the Delta variant, and similarly a Mayo Clinic study of a cohort of patients noted decreased vaccine effectiveness against infection [30] as Delta prevalence increased. By contrast, a cohort study at Kaiser Permanente Southern California [31], in which 47% of infections were classified as Delta, resulted in high (87.4%) effectiveness for the Moderna vaccines.

It is hard to discern a consistent pattern in these studies. Undoubtedly there are demographic, social, and environmental effects that confound simple comparison and interpretation. One observation that could be made is that such studies lack spatial and temporal resolution, and that the ability to follow vaccine effectiveness at a more fine-grained level enables one to shed more light on the question of whether or not the Delta variant reduces vaccine effectiveness. We believe this to be the main accomplishment of this work. We will return to this point in the Discussion, below.

We now describe the data and methods by which we have reached our conclusions.

## Methods

The RT-PCR COVID-19 testing data, broken out by vaccination status, are available from the public sources described in Table 1, although in many cases it is unfortunately necessary to screen-scrape the data from “dashboards” which do not offer the underlying data either as downloadable files or through an API or even from raw image files.

The result is a dataset of locations and date ranges, in which vaccinated and unvaccinated groups of tested individuals, of differing sizes, yield test-positive rates, from which VE^∗^ may be calculated. We consider only positive test rates in the present analysis, so that our VE^∗^ measures effectiveness against infection, as opposed to against severe disease or hospitalization.

We have also downloaded the U.S. SARS-CoV-2 prevalence data from the `Covariants` project [32], which we use to display the rise of the Delta variant relative to others over the period studied. We also aggregated vaccination data from CovidActNow [33] to aid our analyses.

Finally, we have downloaded and transformed data from the CDC on daily vaccination demographics by state [34], in order to attempt to interpret some of our results.

### Bayesian Effectiveness Estimation

For the purpose of estimating local VE^∗^, we adapt the simplified Bayesian method for the analysis of vaccine *efficacy* described in [35]. From the point of view of the method, the distinction between (on the one hand) the treatment and placebo group of a clinical trial, and (on the other hand) differently-sized vaccinated and unvaccinated populations at a place and time is immaterial—the result is a Bayesian posterior density over a risk-reduction parameter, an efficacy in the case of a clinical trial, an effectiveness in the case of uncontrolled case data. The advantage of the procedure is that one may straightforwardly obtain uncertainty bounds— Bayesian credible regions—on the VE^∗^ estimates thus obtained; these uncertainty bounds help give a sense of what spatial and temporal variations in VE^∗^ may or may not be interpreted as mere statistical fluctuations.

To review and adapt the main result of [35]: suppose that the ratio of vaccinated population fraction to the unvaccinated population fraction is *r*; that the number of positive tests among the vaccinated population is *N*_*V*_; and that the number of positive tests among the unvaccinated population is *N*_*U*_. We assume, for simplicity, a uniform prior distribution density *π*(VE^∗^) = 1. Finally, we also assume for simplicity a model in which there is no selection bias that preferentially tests either population more than the other. Then the posterior probability density *π*(VE^∗^|*N*_*V*_, *N*_*U*_) is given by

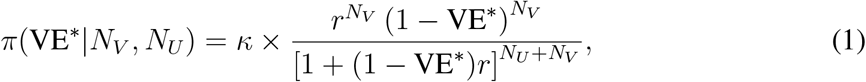

where *κ* is a normalization constant.

One may easily show that if *N*_*V*_ ≠ 0 then the posterior in Equation (1) has a maximum at 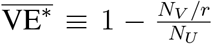, as expected. In addition, the function in Equation (1) may be subjected to a numerical quadrature from which Bayesian credible intervals about the peak VE^∗^ may be obtained. In what follows, whenever we display an error bar for a VE^∗^ in a plot, it corresponds to a 90% Bayesian credible region.

Equation (1) can also be used to *jointly fit* the data from many localities and epochs, in order to obtain a weighted average VE^∗^ over space and time. This can be done by multiplying together the individual posterior densities for each observation, to obtain a “global” posterior density from which a peak and an uncertainty may be extracted. An example of this is shown below, in Figure 1. While a technically-valid procedure, its result should only be interpreted as a convenient weighted average of sorts. The model that this operation represents is one in which all the data really have the same VE^∗^, and the scatter is actually due to statistical noise. As we will presently see, this model does not in fact correctly represent the situation—there are significant variations in VE^∗^ from location to location and over time. Nonetheless, the weighted average allows us to get a sense of the general performance of COVID-19 vaccines during the period of advent of the Delta variant in the U.S.A.

**Figure 1:**
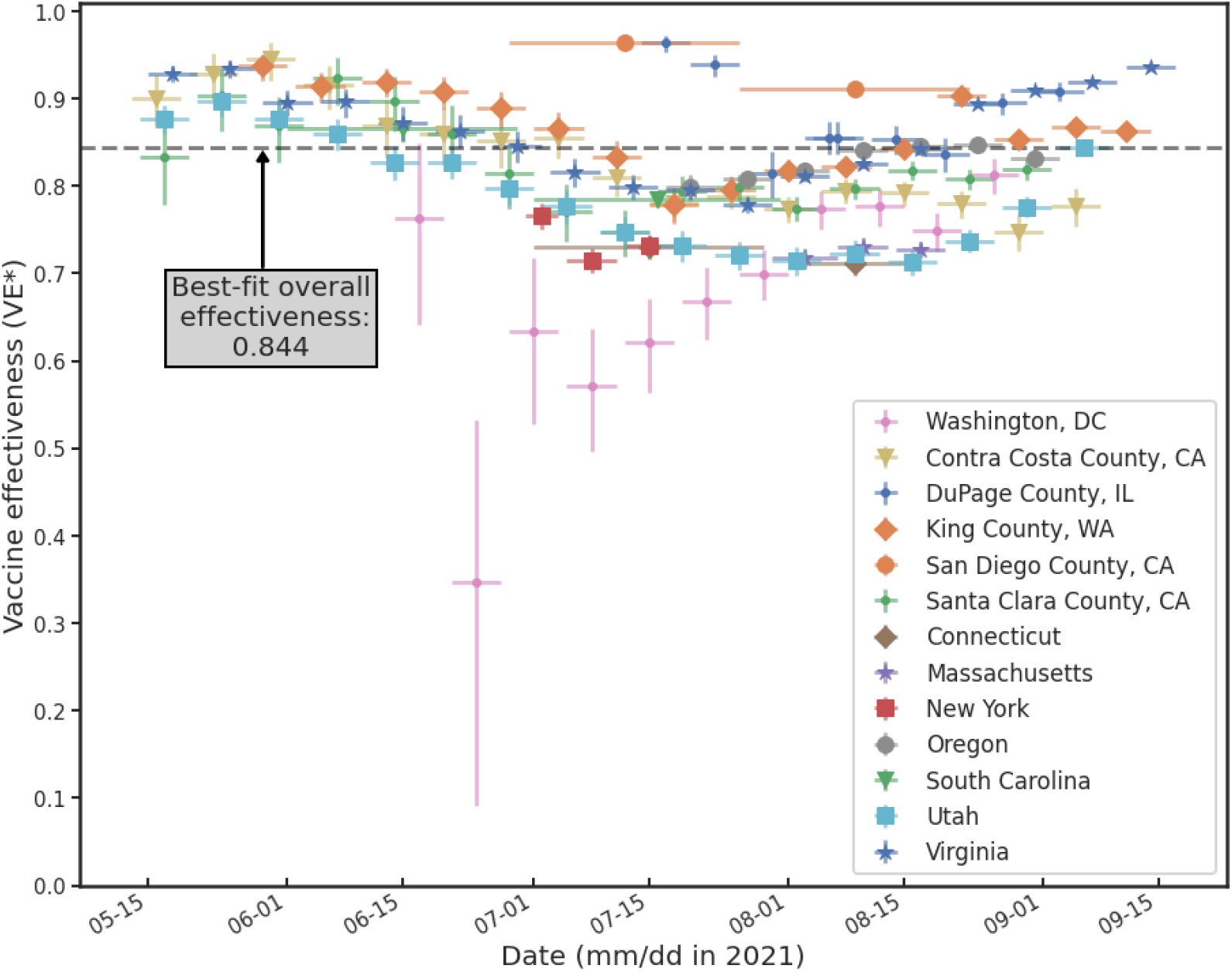
All vaccine effectiveness data, subjected to a joint fit weighted by the Bayesian errors, yields a global effectiveness of 84.1%.

### Interpreting The VE^∗^ Time Series

It is worth discussing the interpretation of a “time-series of VE^∗^.” As we will see below, our estimates of VE^∗^ show evidence of both geographic and temporal variation, with the latter noticeable on timescales as short as a week in some cases. One may well wonder what meaning is to be ascribed to a measure of infection risk reduction that has these kinds of properties.

It is helpful to cast the problem in slightly more formal terms. We can represent VE^∗^ in terms of probabilities as

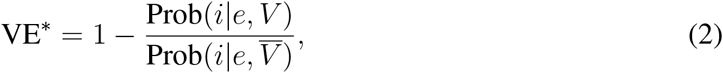

where the symbol *i* represents the proposition “an individual is infected,” *e* represents “the individual was exposed to the virus,” *V* represents “the individual belongs to the vaccinated cohort,” and 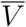 represents the complementary proposition “the individual belongs to the unvaccinated cohort.”

This representation is potentially misleading because of the fraught meanings of *V* and 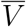, which carry with them not only vaccination status, but also *demographic* information concerning the composition of the respective cohorts. That is, the two cohorts are represented by *distributions* in geography, age, health, race, and many other characteristics. Suppose that these demographic distributions are parametrizable, and that the distributional parameters are denoted by *θ*_*V*_, 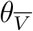 for the vaccinated and unvaccinated cohort, respectively. Then we have a more precise notation to express participation in a particular cohort, to wit

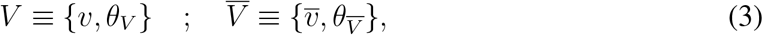

where *v* and 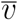 express the non-demographic proposition “*this* individual is (resp., is not) vaccinated”. Now *V* means “this individual (a) is vaccinated, and (b) was drawn from a population with a demographic distribution characterized by the parameters *θ*_*V*_,” and similarly for 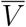. With this interpretation of *V* and 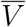, the representation of VE^∗^ in Equation (2) is no longer potentially misleading.

It is now possible to understand how VE^∗^ might be variable in time: it is because *the cohort distribution parameters, θ*_*V*_ (*t*) *and* 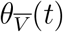 *are subject to temporal variation*. For example, early in the COVID-19 vaccination campaign (i.e., in the early months of 2021) the U.S. followed guidance from the Advisory Committee on Immunization Practices (ACIP), which recommended a phased introduction, with early vaccination phases prioritizing the elderly [36]. This necessarily produced a demographic high age skew in *θ*_*V*_ early on, which declined rapidly in the Spring of 2021, as younger populations began to receive vaccines.

To make the time-dependence evident, we may re-write Equation (2) as follows:

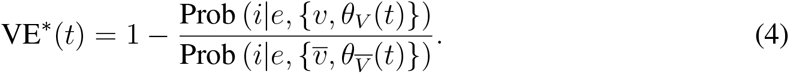

Note that while time-dependent behavior of a risk-reduction factor such as VE^∗^ may seem odd or disquieting, its root cause is the probabilistic conditioning on the demographic information in *V*, 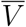, which is usually the same information that epidemiologists have at their disposal and must condition their risk-reduction calculations on. This is a feature, not a bug, of VE^∗^, because in epidemiology, vaccines protect populations, not individuals, and it is demographic information that necessarily conditions questions of risk assessment.

## Results

### Global Behavior of VE^∗^

A global view of all the VE^∗^ fits from all localities at all epochs considered is shown in Figure 1. The most salient feature of the figure is that VE^∗^ remained quite high almost everywhere in the U.S.A. throughout the period from mid-May 2021 to late August 2021. The mean VE^∗^ over all sites and times was 0.841, although from the plot it is clear that there was considerable scatter in the values. In only one location—Washington, DC—did VE^∗^ drop below 0.7, for a period of about a month beginning in late June, and due to small testing volumes the uncertainties in *VE*^∗^ are quite large at this time. Note that the mean VE^∗^ is a *weighted* mean, obtained by a joint fit of the product of the posterior densities in Equation (1). Data with large error bars (that is, corresponding to low test volumes) are naturally given low weight by such a procedure. This explains, for example, why the points from Washington D.C. do not drag the mean value down: those points have large uncertainties, and hence low weight in the fit.

In fact, Figure 1 shows a curious feature that is shared by nearly all locations, which is a slight drop in VE^∗^ between early and mid-July, followed by a recovery of VE^∗^ by early September. Since the vaccinated and unvaccinated populations are not controlled in these studies, the behavior in these time series almost certainly reflects demographic shifts in the constitution of the two groups. We will examine one such shift—average age of the vaccinated group—below.

In Figure 2, we show time-resolved joint fits of VE^∗^ over all localities when using seven-day (left) and 14-day (right) data bins. We se again the mid-summer dip, although with an anomalous high point around 15 July. Comparison with Figure 1 suggests that this anomaly is due to the inhomogeneity of the distribution, with the VE^∗^ due to San Diego county strongly “dissenting” from VE^∗^ due to the rest of the U.S.A. The remaining data in Figure 1 appears to move together more harmoniously, suggesting that the average behavior displayed in Figure 2 is robust and informative of a national coherence in vaccine demographic trends. By both the seven-day and 14-day bin time series, measured VE^∗^ started out at a value of about 0.9 in mid-May, declined slightly to about 0.76 by mid-July, only to recover to a value of about 0.9 by early September.

**Figure 2:**
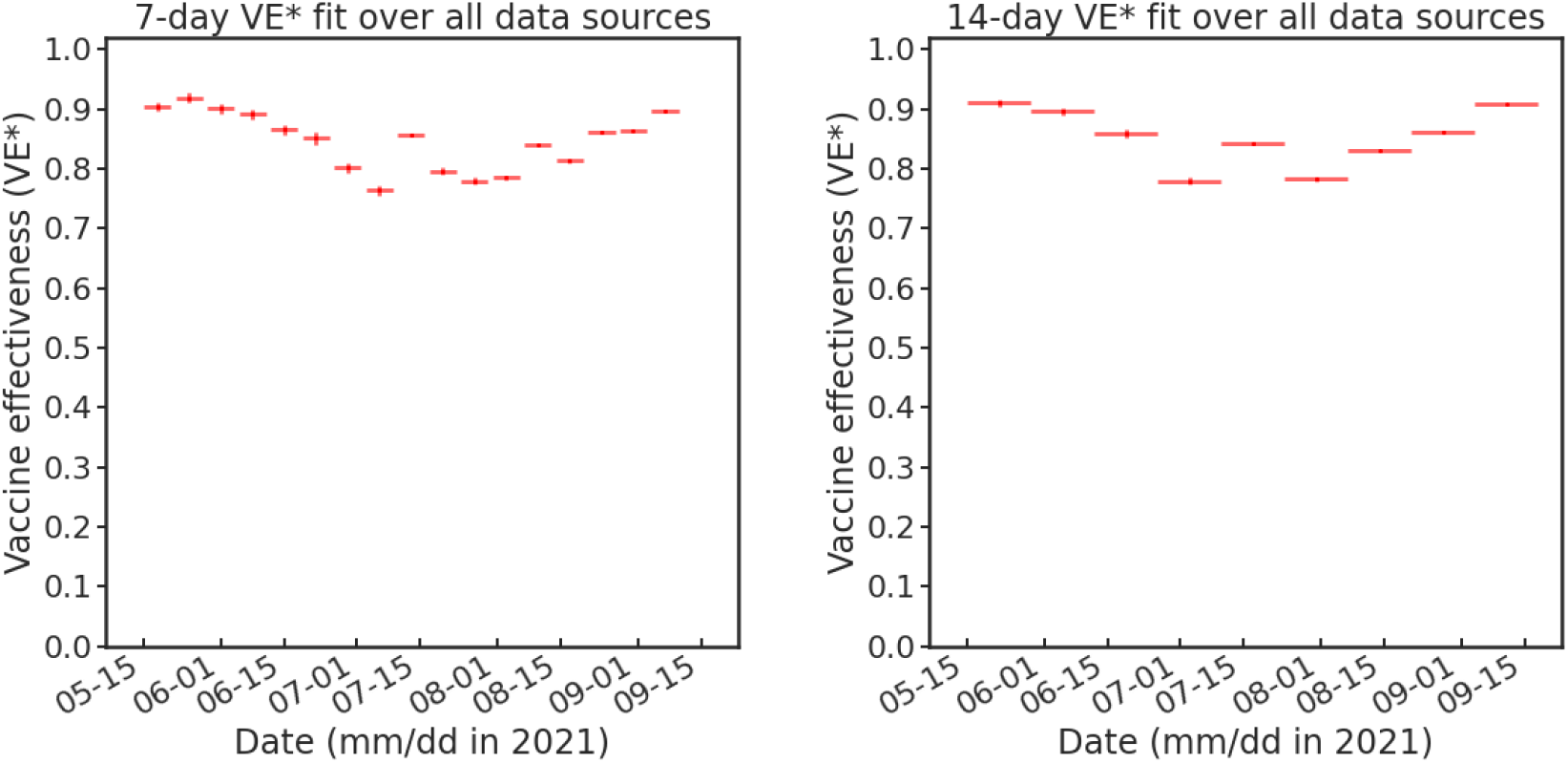
Time series of weekly VE fits to data from all available sites. Left panel: Seven-day averages. Right panel: 14-day averages. Note the recovery of VE to a value of about 0.9 by early September 2021.

### Local Behavior: VE^∗^ Versus Delta

Figure 3 displays the VE^∗^ fit data for the five individual U.S. counties plus Washington, DC, for which we obtained test-positive data by vaccination status. The plots also show the progress of the Delta (B.1.617.2) variant of SARS-CoV-2, which as can be seen from the upper histograms rapidly ascended from essentially negligible prevalence to dominance of the viral genome in the space of the three months of the study. What is apparent from these figures is the contrast between the dramatic advent of the Delta variant and the relatively modest changes in VE^∗^ that were happening concurrently in every region. It is evident from these figures that to the extent that the Delta variant had an impact on VE^∗^, that impact was of minor importance. Had the impact of Delta on VE^∗^ been a dominant, or even an important, effect, the data would show a monotonic decrease in VE^∗^ corresponding to and correlated with the monotonic increase of Delta prevalence. Instead the data shows VE^∗^ fluctuations, some rising, some falling, some even oscillating, and these are presumably driven by demographic and environmental factors affecting vaccination statistics, and which are clearly overwhelming any effect on VE^∗^ that may be due to the rapid ascent of Delta prevalence.

**Figure 3:**
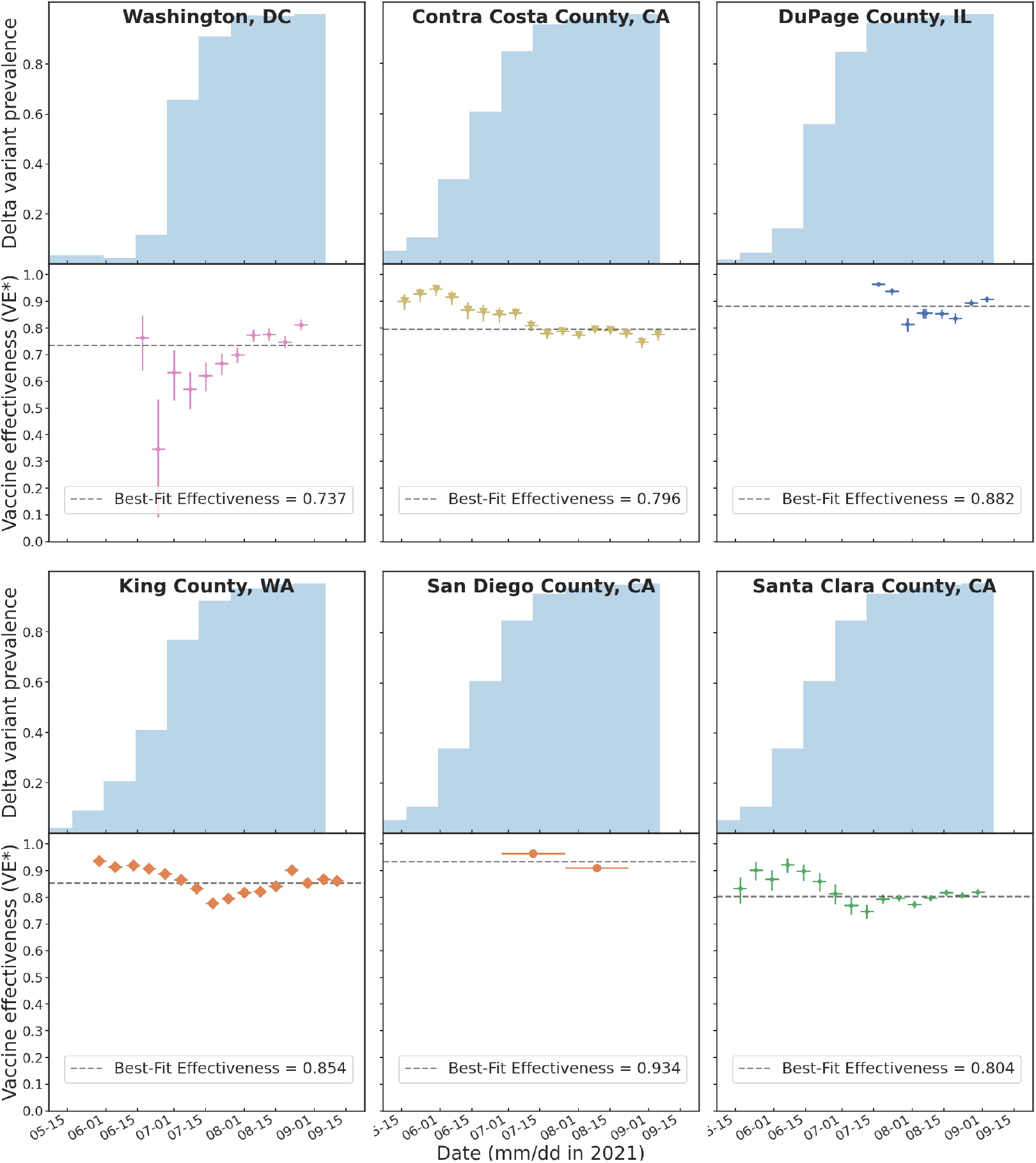
In Washington DC, and five counties for which data are available, vaccine effectiveness was quite stable and high, while Delta prevalence grew from essentially zero to nearly 100% in three months.

Figure 4 displays the same plots but for state-level data for the seven U.S. states for which we were able to obtain the necessary data. The data from the states tells the same story as the county-level data: there is no trend of systematically dropping VE^∗^ such as would be expected if the Delta variant were capable of strongly evading vaccine-stimulated immune response. We conclude from these data that there is no evidence that the Delta variant escapes immunity from the COVID-19 vaccines in use in the U.S.A., and that there is therefore no evidence that the variant causes additional breakthrough infections, above and beyond the infections already expected due to the known imperfect effectiveness and efficacy of the available COVID-19 vaccines.

**Figure 4:**
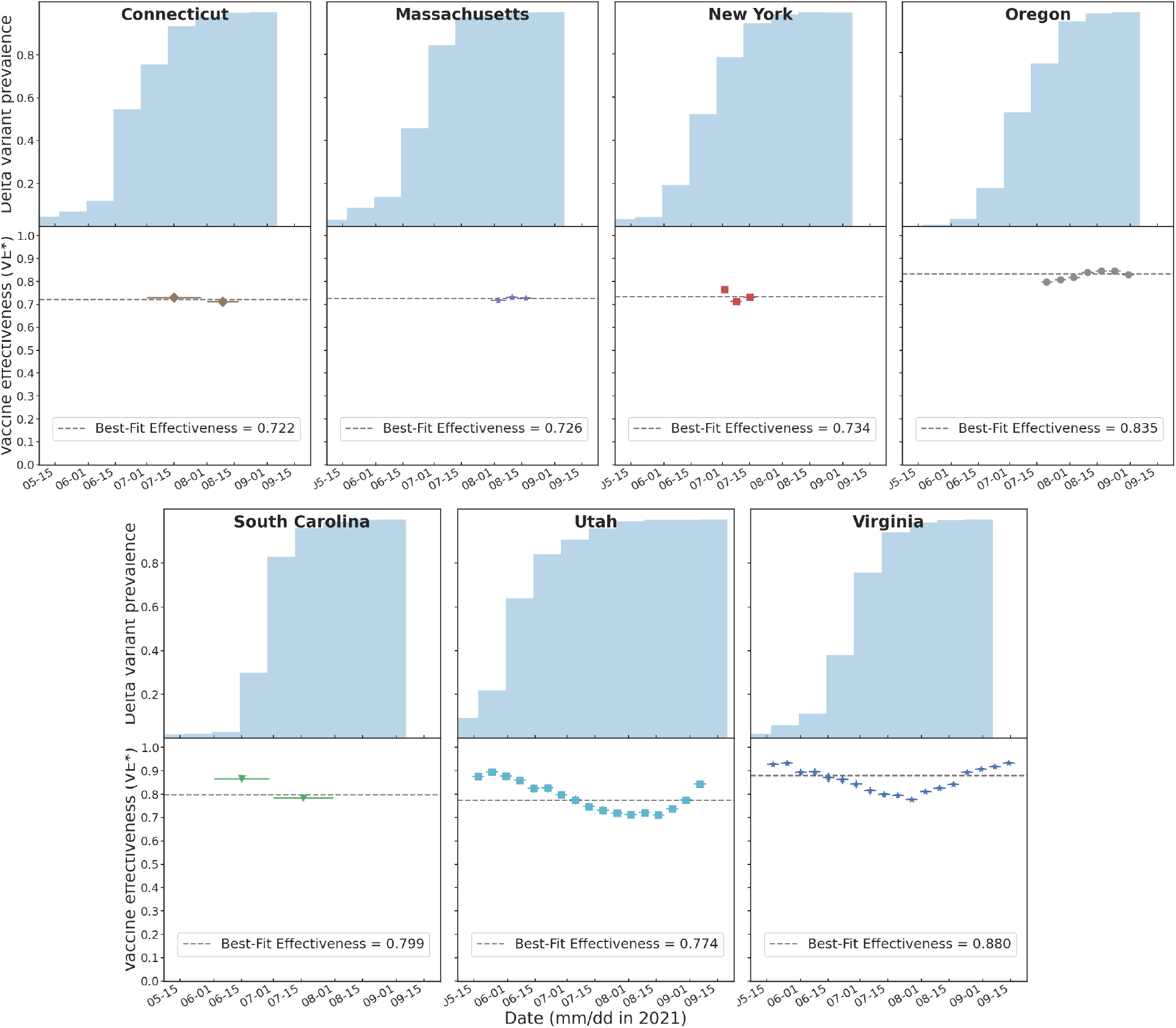
In U.S. states, there are fewer available data, but the behavior is similar to that of the counties.

### Vaccine Demographics

The modulation of the VE^∗^ time series in Figures 1–4 is a noteworthy feature of the data. We spent some effort speculating about its origin, and investigating possible other data correlations that might explain it. Given the relatively fragmented and unsystematic state of present epidemic surveillance data infrastructure, it is not easy to investigate many possibilities. We noticed that a CDC dataset [34] makes it possible to crudely bin the vaccinated population into bins of ages 12–18, ages 18–65, and ages 65+. From these age bins, it is possible to construct a rough-and-ready measure of mean age of the vaccinated population. Since the dataset contains daily statistics by state, it is possible to view, on a state-by-state basis, the average age of the vaccinated cohort in the U.S.A., albeit according to a rather crude measure of “average age.” This is an example of a time-dependent demographic distributional parameter in the set *θ*_*V*_ (*t*), discussed above in the subsection of the Introduction on interpretation of time-series of VE^∗^.

The time series of average age of the vaccinated cohort for the U.S. states that concern the present study are displayed in Figure 5. We see that in every state studied, the mean age of the vaccinated cohort dropped significantly over the summer of 2021, in many cases by as much as four years over three months. This rapid drop in mean age of the cohort was undoubtedly caused by the approval on 10 May 2021 by the US Food and Drug Administration of Emergency Use Authorization (EUA) for adolescents aged 12-15 of the Pfizer/BioNTech COVID-19 vaccine [37]. During this time, younger and, on average, presumably healthier people left the unvaccinated cohort and entered the vaccinated cohort, which one would expect, in principle, to raise VE^∗^. Possibly this effect may have had something to do with the “recovery” of VE^∗^ following the July 15 minimum. However, the root cause of the minimum and subsequent recovery is still a puzzle. Such changes in VE^∗^ may be consistent with changes in the vaccinated and unvaccinated cohort demographics, changes in social behavior, or environmental or seasonal effects, each of which is worth independent analysis and investigation.

**Figure 5:**
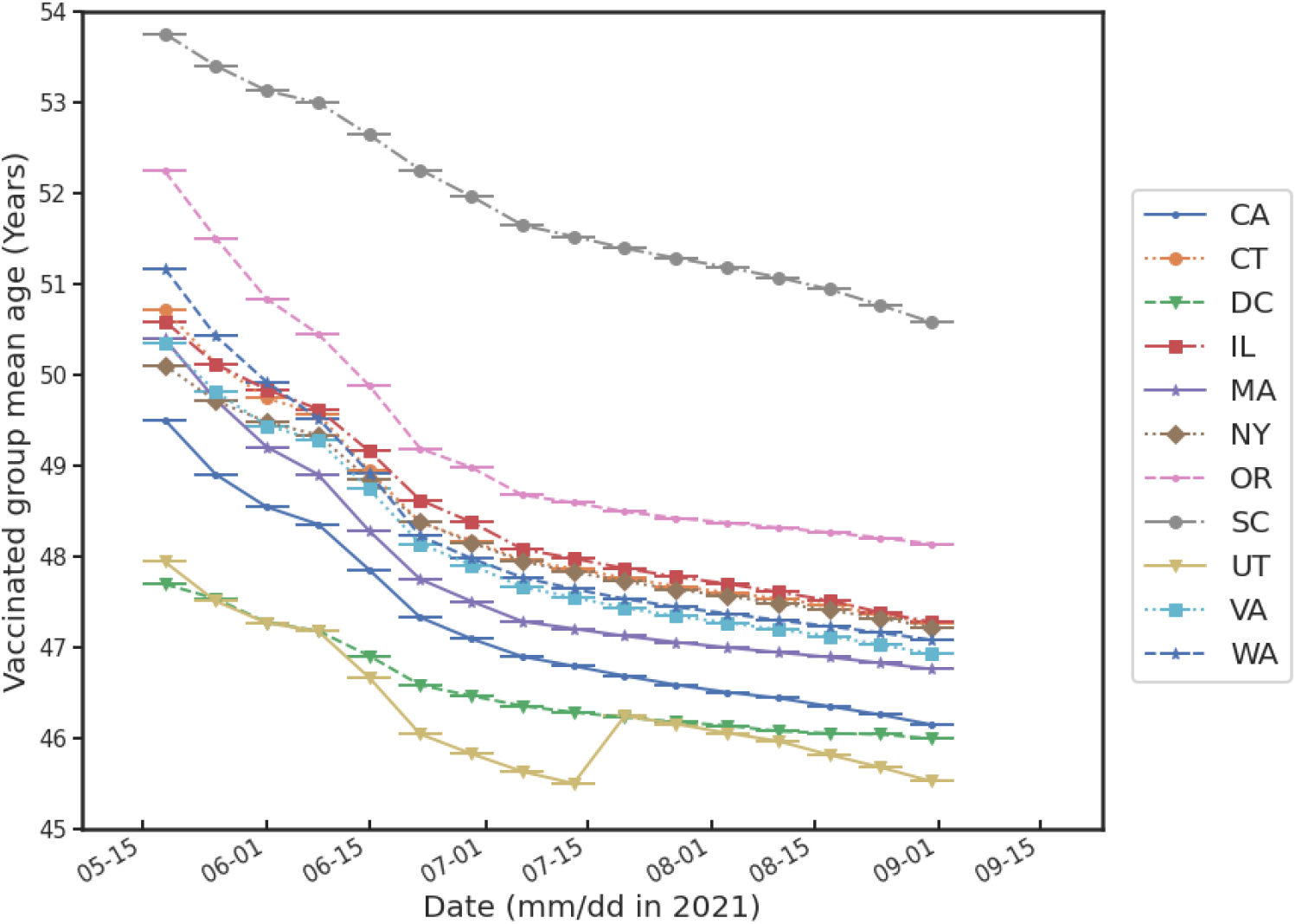
Mean age of vaccinated groups dropped markedly in every U.S. state between May and August 2021.

## Discussion

The behavior of VE^∗^ between 15 May 2021 and 15 September 2021 may be broadly characterized as a nationwide gentle decline from an initial set of values clustered around 0.9 on 15 May 2021, to a minimum with a mid-July average of around 0.76, followed by a recovery to values clustered once again around 0.9 by early September. Various sites around the U.S.A. moved broadly in harmony with this trend, with discrepancies from the trend presumably representing local idiosyncrasies in the demographics of vaccine distribution. The “wave” behavior of VE^∗^ seems potentially interpretable as a transfer of demographic risk factors such as age, comorbidities, and other clinical vulnerabilities, between the vaccinated and unvaccinated cohorts, as vaccination rates increased in various demographic groups. But despite these variations, it is clear that the rapid growth of the Delta variant in the U.S.A., from near-zero prevalence to near-dominance of the SARS-CoV-2 genome in the three months between 15 May 2021 and 15 August 2021, has had a negligible impact on VE^∗^.

The clarity of the outlook afforded by the analysis presented here stands in contrast to the murky picture regarding the effect of Delta on vaccine-induced immunity to COVID-19 that emerged from the literature [25, 26, 27, 28, 29, 30, 31] reviewed in the Introduction. We suggest that the difference is that those analyses were necessarily coarse-grained, representing a snapshot (or a comparison of two snapshots) in time, and usually aggregating data into a single geographic unit, even in the case of widely-scattered studies such as [27, 29]. In contrast, and particularly in light of the interpretation of VE^∗^ time-series discussed above, the present study builds on a *geographically- and temporally-resolved view of VE*^∗^, *interpreted as an actionable risk-reduction factor, with error bars, and explicitly probabilistically conditioned on local, real-time vaccine demographics*.

The power of this type of analysis, which can be seen as a contribution to the growing field of “outbreak analytics” [38, 39], is due in part to the simplicity of the data necessary to drive it (COVID-19 RT-PCR data, separated by vaccination status); in part to the informativeness of the time-series of VE^∗^ that are output (as illustrated by what could be learned by comparing them to the time-series of Delta prevalence); and in part to the fact that VE^∗^, as estimated here, is precisely the demographic risk-reduction estimate that is of interest from an epidemiological point of view: an epidemiologist would want to know, for example, “how much better was the vaccinated *population* of DuPage County, IL, protected against COVID-19 than their unvaccinated peers on September 1 2021?” This kind of question is what our VE^∗^ estimates can answer.

Given these observations on the method presented in this article, we see great potential for transforming at least that aspect of epidemic surveillance that pertains to monitoring the effectiveness of COVID-19 vaccines in the face of the inevitable emergence of future new strains of SARS-CoV-2, both in the U.S.A and internationally. What is required for this to happen is some reform in the data-management and publication practices of public health authorities with respect to the availability of COVID-19 RT-PCR test data sorted by vaccination status.

In the first place, it would clearly be beneficial if *all* public health organizations that aggregate COVID-19 test data would gather vaccination status with each test, if they do not do so already, and publish their test-positive statistics broken out by vaccination status. Additionally, it would be helpful if *testing volume* data, i.e., the total number of tests performed for each site, were also broken out by vaccination status. This would permit checking (and generalizing) the model assumption that both the vaccinated and unvaccinated groups are tested at equal rates, which at the moment is unvalidated in our method.

Our experience in gathering such data suggests several opportunities for public health departments to improve their data reporting. First, data should be made available in open machine-readable formats, e.g., comma separated value (CSV) files, embedded Javascript object notation (JSON), or via programmatic interfaces such as REST APIs [40] to facilitate automated collection. Second, the data should be made available in formats and structures that are common with those used by other such departments. (Open source software can facilitate such sharing of approaches.) These first two improvements could be achieved, for example, by making vaccine effectiveness data available via simple export from a dashboard as a CSV, via download of the underlying raw data files (e.g., as done by Utah), or by embedding JSON documents in the HTML using a format such as the Schema.org Dataset type [41] used for Google Datasets. Third, it may be important to create a central registry of such data that would enable researchers to easily discover the existence of such resources, and would enable searching, querying, and browsing by the geography, data types, and more. By realizing each of these improvements, the future collection and utilization of these critical data by researchers would become much simpler.

We note several potential sources of bias in the datasets analyzed in this work that may limit the effectiveness of the methodology described here. 1) We have observed retrospective data revisions in the collected data streams. These revisions change the calculated values, and necessitate recurring data collection and analysis methods. 2) The data streams that we have collected may not be fully representative of the nation as a whole. We found the datasets analyzed in this study by performing Google searches with key phrases (e.g. “Utah by vaccination status”) and by monitoring and searching Twitter for mention of locations reporting cases or population risk by vaccination status. This methodology was inherently ad hoc, and has assuredly missed some data sources. To address this, a future data infrastructure should allow for simple addition of both new data streams and the scrapers needed to collect their data from their source. 3) Many of the data analyzed in this work were made available through the web in formats that are not conducive to automated extraction of the key values (e.g., as images, within natural language text, or via dashboards without export functionality). The processes used to collect such data (e.g., transcribing values by hand) are prone to human error. 4) Data presented on public websites are often available only temporarily, necessitating a snapshot, and precluding retrospective analyses (even using the Internet Archive, which caches website HTML and some images, but not dashboard content). These and other issues would be remedied if data were made available in machine-readable formats and if versioning was applied to the data files for each data stream.

## Data Availability

All data sources are provided and described in the publicly available GitHub repository referenced in the manuscript.

https://doi.org/10.26311/CF4K-VM12

## Acknowledgements

This material is based upon work supported by the U.S. Department of Energy, Office of Science, under contract number DE-AC02-06CH11357.

## Government License

The submitted manuscript has been created by UChicago Argonne, LLC, Operator of Argonne National Laboratory (“Argonne”). Argonne, a U.S. Department of Energy Office of Science laboratory, is operated under Contract No. DE-AC02-06CH11357. The U.S. Government retains for itself, and others acting on its behalf, a paid-up nonexclusive, irrevocable worldwide license in said article to reproduce, prepare derivative works, distribute copies to the public, and perform publicly and display publicly, by or on behalf of the Government. The Department of Energy will provide public access to these results of federally sponsored research in accordance with the DOE Public Access Plan. http://energy.gov/downloads/doe-public-access-plan.

